# COVID-19: Effectiveness of Non-Pharmaceutical Interventions in the United States before Phased Removal of Social Distancing Protections Varies by Region

**DOI:** 10.1101/2020.08.18.20177600

**Authors:** William K. Pan, Stefanos Tyrovolas, Giné-Vázquez Iago, Rishav Raj Dasgupta, Fernández Daniel, Ben Zaitchik, Paul M. Lantos, Christopher W. Woods

## Abstract

Although coronavirus disease 2019 (COVID-19) emerged in January 2020, there is no quantified effect size for non-pharmaceutical interventions (NPI) to control the outbreak in the continental US. Objective. To quantify national and sub-national effect sizes of NPIs in the US. Design. This is an observational study for which we obtained daily county level COVID-19 cases and deaths from January 22, 2020 through the phased removal of social distancing protections. A stepped-wedge cluster-randomized trial (SW-CRT) analytical approach is used, leveraging the phased implementation of policies. Data include 3142 counties from all 50 US states and the District of Columbia. Exposures. County-level NPIs were obtained from online county and state policy databases, then classified into four intervention levels: Level 1 (low) – declaration of a State of Emergency; Level 2 (moderate) – school closures, restricting nursing home access, or closing restaurants and bars; Level 3 (high) – non-essential business closures, suspending non-violent arrests, suspending elective medical procedures, suspending evictions, or restricting mass gatherings of at least 10 people; and Level 4 (aggressive) – sheltering in place / stay-at-home, public mask requirements, or travel restrictions. Additional county-level data were obtained to record racial (Black, Hispanic), economic (educational level, poverty), demographic (rural/urban) and climate factors (temperature, specific humidity, solar radiation). Main Outcomes. The primary outcomes are rates of COVID-19 cases, deaths and case doubling times. NPI effects are measured separately for nine US Census Region (Pacific, Mountain, West North Central, East North Central, West South Central, East South Central, South Atlantic, Middle Atlantic, New England). Results. Aggressive NPIs (level 4) significantly reduced COVID-19 case and death rates in all US Census Regions, with effect sizes ranging from 4.1% to 25.7% and 5.5% to 25.5%, respectively, for each day they were active. No other intervention level achieved significance across all US Regions. Intervention levels 3 and 4 both increased COVID-19 doubling times, with effects peaking at 25 and 40 days after initiation of each policy, respectively. The effectiveness of level 3 NPIs varied, reducing case rates in all regions except North Central states, but associated with significantly higher death rates in all regions except Pacific states. Intervention levels 1 and 2 did not indicate any effect on COVID-19 propagation and, in some regions, these interventions were associated with increased COVID-19 cases and deaths. Heterogeneity of NPI effects are associated with racial composition, poverty, urban-rural environment, and climate factors. Conclusion. Aggressive NPIs are effective tools to reduce COVID-19 propagation and mortality. Reducing social and environmental disparities may improve NPI effects in regions where less strict policies are in place.

## Introduction

COVID-19 (novel coronavirus disease-2019) has spread rapidly across the globe, infecting over 20 million people with the SARS-CoV-2 virus. One-fourth of all reported cases and deaths have occurred in the United States. Public health interventions to reduce spread of COVID-19 in the US have varied in timing and types of interventions undertaken. No official US policy existed to prevent COVID-19 transmission until January 31, 2020, when a presidential order blocked entry of non-US citizens into the US traveling from China. The state of Washington declared the first State of Emergency (February 29), followed by California (March 4) and Maryland (March 5); however, many counties implemented restrictive policies prior to state action, for example: three counties in Washington (King, Pierce, Snohomish) and four in Arkansas (Grant, Jefferson, Pulaski, Saline) ordered school closings on March 12 vs. statewide closures on March 17; counties in Pennsylvania, California, and New Jersey closed non-essential businesses prior to the state; and counties in California and Idaho issued restrictions on mass gatherings before state policies.

Reasons for early action by counties varies according to their ability to legislate a response as well as socio-environmental issues that influence local disease risk. Such issues include racial disparities, with a disproportionate number of Black and Hispanic Americans reported to be infected or dying [1–3], urban vs. rural characteristics that influence transmission and policy implementation [4], and disease spillover from neighboring counties, which can be exacerbated by economic disparities and shared environmental risks [5]. Environmental factors hypothesized to influence COVID-19 transmission include relative humidity (RH), temperature and UV exposure; however, studies vary, with some finding no effect [6], inverse relationships [7], or mixed effects [8, 9]. Elevated humidity has been associated with an increase in organic aerosols and higher levels of small particulate air pollutants (PM2.5) [10, 11], which have been correlated with transmission [12].

Non-pharmaceutical interventions (NPI) are needed to control COVID-19. Unfortunately, without a randomized trial to quantify the effect size of NPIs or the potential causes of heterogeneity across different regions of the US, policy-makers have relied on modeling studies and early evidence from Asia to guide decisions. For example, susceptible-exposed-infectious-recovered (SEIR) models have attempted to quantify the effects of isolation and contact tracing [13] and to predict how NPI effectiveness influences demand for critical care resources [14]. Studies in Wuhan and Hong Kong have reported how strict interventions (i.e., quarantine, social distancing, shelter in place and active case detection) reduced the COVID-19 reproductive number (R_0_) [15, 16]. Understanding heterogeneity across the US has been particularly challenging. The Institute for Health Metrics and Evaluation (IHME) uses mean relationships from 13 communities to develop prior distributions of the effect of NPIs in their COVID-19 forecasts [17], while a recent analysis of NPIs in the US estimated potential COVID-19 rebound if interventions ended for the country as a whole, but not regionally [18].

The goal of this study is to evaluate national and sub-national effects of NPIs in the US from implementation to Phase 1 reopening by state governments (if specified, otherwise to May 29). In addition, we evaluate potential socio-environmental policy mediators. We leverage the phased implementation of policies at the county and state levels, using a stepped wedge cluster-randomized trial (SW-CRT) analytical approach [19]. We report NPI effects on COVID-19 daily case incidence, doubling time, and reported deaths across nine US Census Regions.

## Methods

### Outcome data

County-level daily cases and deaths of COVID-19 were obtained from all 50 US states and the District of Columbia from January 22 through May 29, 2020. Data are primarily from the Johns Hopkins University Center for System Science and Engineering Coronavirus Resource Center (JHU-CSSE, https://coronavirus.jhu.edu/); however, we evaluated the time series for all counties, correcting any instances when cumulative case or death counts declined over time or when large discrepancies existed with cases or deaths from county and state health department websites (where daily data are available).

### Policy data

We used online policy databases to record the effective date of each public health intervention and the phased reopening at the state and county level [20–24]. Missing county policies were obtained by searching the county’s state government website or conducting a systematic search of grey literature for each county’s policies. We categorized 12 policies into 4 levels of disease control following the New Zealand alert system and Oxford classification [25, 26]: Level 1 (low) – governor declaration of a State of Emergency; Level 2 (moderate) – school closures, restricting access (visits) to nursing homes, or closing restaurants and bars; Level 3 (high) – non-essential business closures, suspending non-violent arrests, suspending elective medical procedures, suspending evictions, or restricting mass gatherings of at least 10 people; and Level 4 (aggressive) – sheltering in place / stay-at-home, public mask requirements, or travel restrictions. The two federal policies blocking entry to the US for non-US citizens (i.e., from China issued January 31 and from Schengen European countries issued March 11) had no effect on COVID-19 morbidity or mortality propagation; thus we classified them as the “non-intervention” period. Phase 1 reopening was defined as opening non-essential businesses (with capacity restrictions), allowing public gatherings of more than 10 people, opening public spaces, or easing shelter in place orders. NPI effects were measured up to five days after Phase 1 reopening or through May 29.

### Demographic and Environmental Data

County level demographic data are from the 2018 American Community Survey [27]. This includes age, sex and racial composition, migration, and educational data, population density (1000 people per square-km) [28], and poverty [29]. The USDA Rural-Urban Continuum Code was used to categorize counties into nine levels of rural-urban characteristics [30]. Three levels indicate metropolitan areas of (1) 1 million or more people, (2) 250,000 to 1 million people, and (3) fewer than 250,000 people. Four urban levels classified by size and adjacency to a metropolitan area: (4) 20,000 or more people adjacent; (5) 20,000 or more, not adjacent; (6) 2,500 to 19,999, adjacent; and (7) 2,500 to 19,999, not adjacent. And two rural levels: (8) less than 2,500 population, adjacent to a metropolitan area; and (9) less than 2,500 population, not adjacent. Environmental data are from the North American Land Data Assimilation System (NLDAS). The NLDAS provides several daily hydrometeorological measures, for which we define 10-day temporal lags of minimum air temperature (Celsius), specific humidity (g/kg) and bias-corrected shortwave radiation (W/m^2^).

### Statistical Methods

Outcomes were defined at the county level as the number of new daily cases, new deaths, and case doubling time. Doubling time is the number of days required to double the cumulative case count on a particular day. Policy levels were time-varying, coded as a 1 while a level was active and 0 otherwise, and as the number of days since an intervention level was initiated. Analyses were stratified by the nine US Census Regions (Pacific, Mountain, West North Central, East North Central, West South Central, East South Central, South Atlantic, Middle Atlantic, New England) to evaluate differential policy effects.

To evaluate intervention levels and socio-environmental factors, the Hussey and Hughes SW-CRT framework was used [31] that takes advantage of the phased county implementation to compare incidence rates within and between counties (see Supplement for details). We use negative binomial mixed models to fit this model, adjusting for period (time series day, beginning January 22), US Census Region, intervention level, and a nested county*state random effect. The model for cases and deaths included total county population as an offset. Model selection was conducted with AIC. Variables considered include: rural-urban continuum code; minority (Black and Hispanic) and total population density; net county migration rate in 2018; percent of the 2018 county population Black (alone or mixed race), Hispanic (alone or mixed race), living in poverty, or with a college education or higher; and climate parameters (10-day lags for minimum temperature, specific humidity, and UV radiation). Models for cases and deaths were run separately for each US Census region (combining east and west North Central, and east and west South Central states), while doubling time included interactions to evaluate regional heterogeneity. Final models were fit using the R package glmmTMB (R V3.6.3)[32] and SAS 9.4 with Gaussian adaptive quadrature.

## RESULTS

Among the 3142 counties from which we obtained data, 339 (10.8%) from 26 states created policies prior to their state government, the majority (211 or 62%) were located in Texas, Nebraska, Missouri, and Pennsylvania. Counties initiating policies prior to the state were more likely to be located in metropolitan areas (17.5% metro counties vs. 6.8% non-metro countries adopted early policies, p< 0.0001), have populations with higher educational levels, higher percentages of Hispanic population, and fewer people living in poverty (respectively, counties with early policy adoption had 26.3%, 15.9%, and 13.8% of their populations with a college degree, Hispanic descent [mixed or alone], and living in poverty, vs. 21.0%, 8.5%, and 15.3% for countries that did not adopt early policies, p< 0.0001). The time between the first COVID-19 case and the initiation of any policies varied, with coastal regions (Pacific, Mountain, New England, and Middle Atlantic) having longer delays compared to central regions (Figure 1). However, middle America regions also benefited from the experience of coastal regions as the first COVID-19 cases occurred an average of 10 days prior in coastal vs. inland regions, providing critical time for NPI vetting and initiation. Once initiated, policy duration averaged 5.7, 3.6, 11.9 and 44.3 days for levels 1-4, respectively, with significant variation by state and some states having zero days for any particular level (Supplemental Table A1). Note that county and state governments in New England and Mountain states were more likely to initiate policies before the first reported COVID-19 death compared to other regions (data not shown).

**Figure 1.**
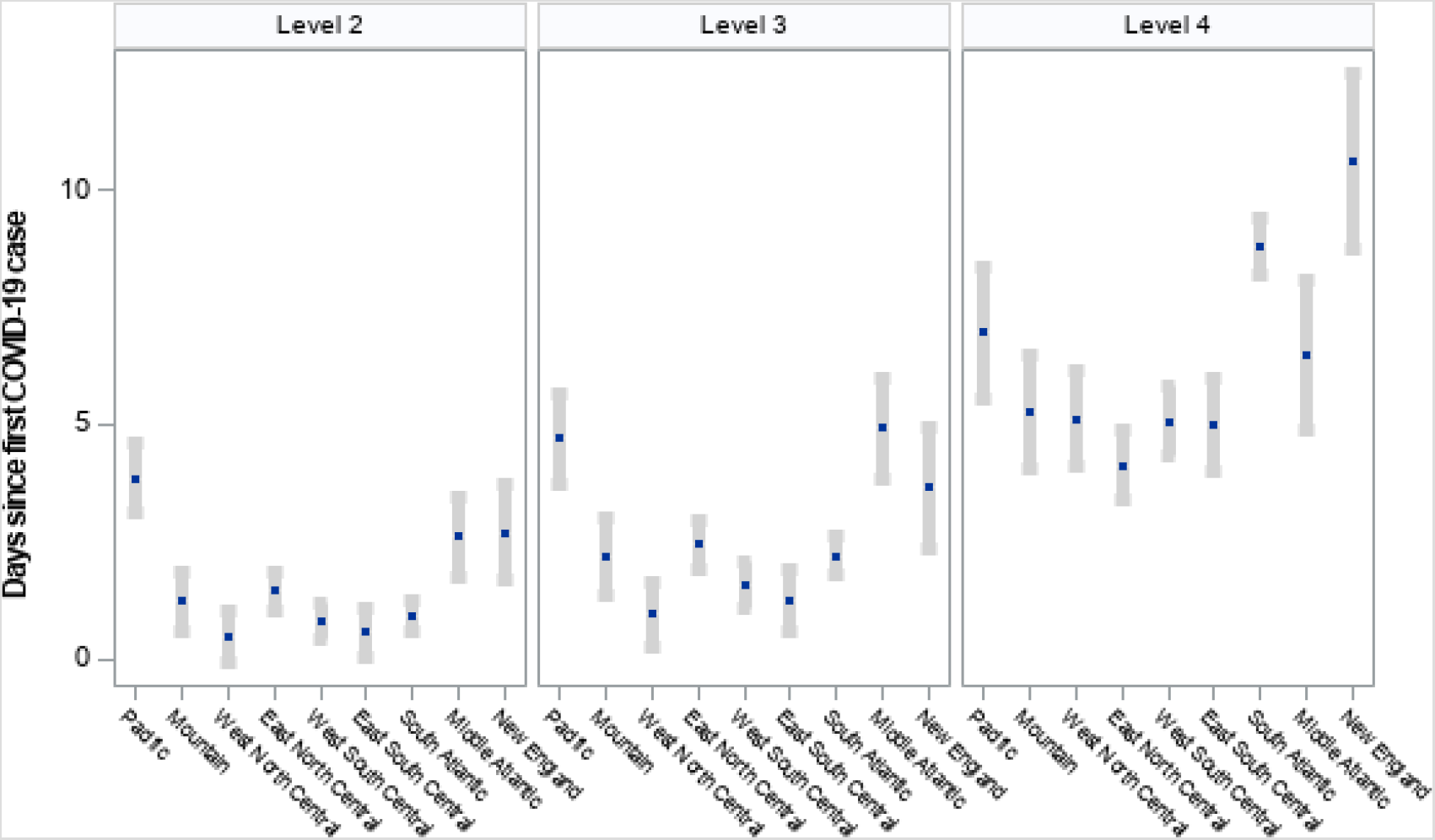
Time between first COVID-19 case to Policy Initiation by US Census Region (mean days and 95% confidence interval) ^1^Policies classified under each level of intervention: Level 2—school closures, restricting access to nursing homes, closing restaurants and bars; Level 3—non-essential business closures, suspending non-violent arrests, suspending elective medical procedures, suspending evictions, restricting mass gatherings of at least 10 people; Level 4—sheltering in place / stay-at-home, mask requirements in public, travel restrictions ^2^States classified for each region are as follows: Pacific—CA, WA, OR, HI, AK; Mountain—MT, WY, ID, NV, UT, CO, AZ, NM; West North Central—ND, SD, NE, KS, MN, IA, MO; East North Central—WI, IL, MI, IN, OH; West South Central—TX, OK, AR, LA; East South Central—KY, TN, MS, AL; South Atlantic—FL, GA, SC, NC, VA, WV, DC, MD, DE; Middle Atlantic—NY, PA, NJ; New England—ME, CT, NH, MA, CT, RI

### Policy Effects on COVID Propagation and Mortality

#### Case Rates

Model results indicate that intervention level 4 achieved at least a 50% reduction in COVID-19 case rates in six days (95% CI for case reduction in six days: 36.0%-61.8%), compared to eight days for intervention level 3 (95% CI: 33.8%-68.4%, Figure 2, Table 1). Intervention levels 1 and 2 never achieved a 50% reduction; in fact, level 1 policies resulted in a 50% increase in cases within 4 days (95% CI for case increase: 14%-123%) and, if level 2 policies remained in place, a 50% increase in cases may have occurred in 34 days (95% CI for case increase: 30%-730%).

**Figure 2.**
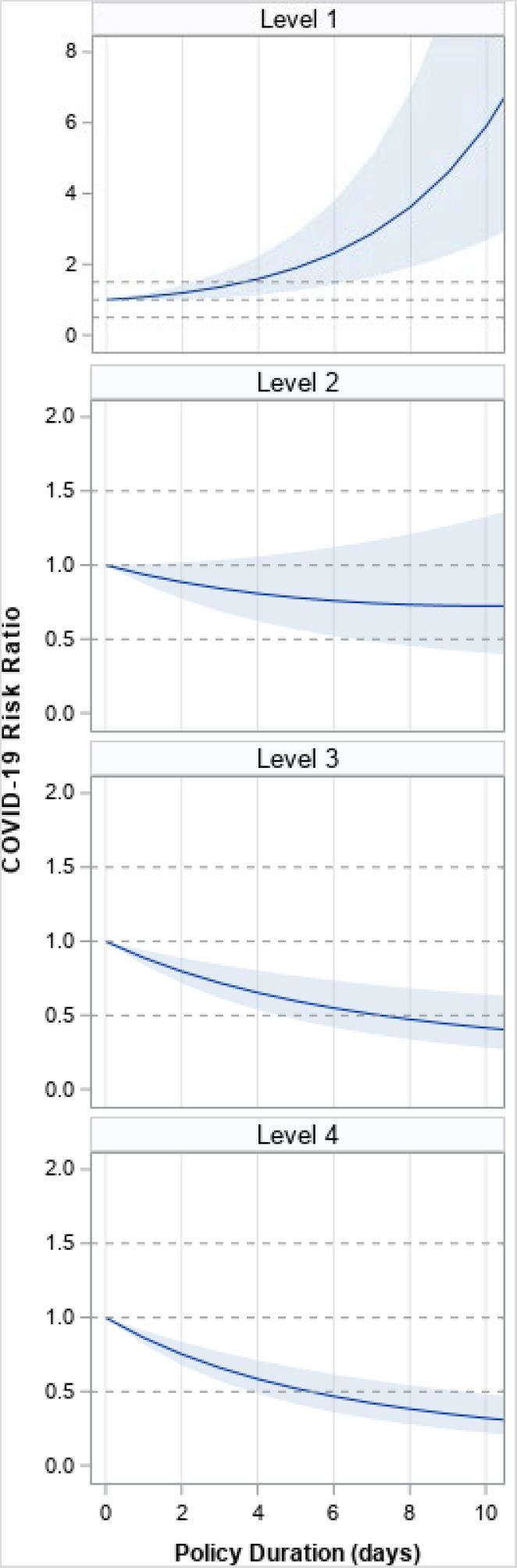
Mean policy effect on COVID-19 case rate during the first 10 days after policy initiation. Mean effect sizes for intervention level duration (solid blue line) with 95% confidence intervals (shaded blue). Dotted lines represented reference values for no change (1), 50% increase (1.5) and 50% decrease (0.5). Effect sizes are from negative binomial models as described in methods that adjust for: period; rural-urban continuum classification; 2018 county population percentage with at least a bachelor’s degree, living in poverty, Black, and Hispanic; and 2018 net county migration rate. Effect sizes are a weighted mean across US Regions, weights defined as the number of counties per region. Full regression results shown in Supplemental Table B1.

**Table 1.**
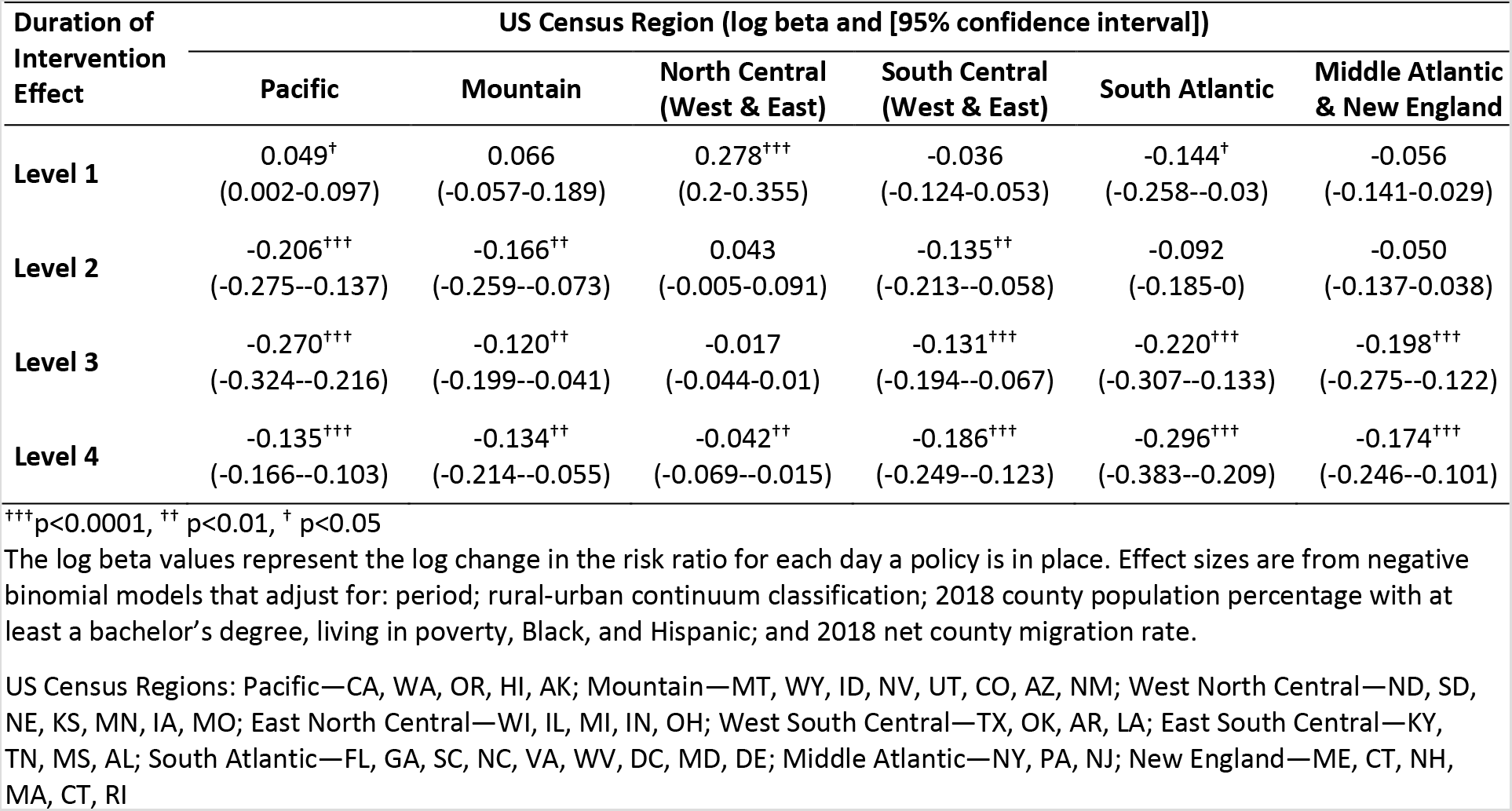
Change in Log Risk Ratio for each day an intervention is in effect by US Census Region.

| Duration of Intervention Effect | US Census Region (log beta and [95% confidence interval]) |  |  |  |  |  |
| --- | --- | --- | --- | --- | --- | --- |
|  | Pacific | Mountain | North Central (West & East) | South Central (West & East) | South Atlantic | Middle Atlantic & New England |
| **Level 1** | 0.049<sup>†</sup><br>(0.002-0.097) | 0.066<br>(-0.057-0.189) | 0.278<sup>+++</sup><br>(0.2-0.355) | -0.036<br>(-0.124-0.053) | -0.144<sup>†</sup><br>(-0.258--0.03) | -0.056<br>(-0.141-0.029) |
| **Level 2** | -0.206<sup>+++</sup><br>(-0.275--0.137) | -0.166<sup>++</sup><br>(-0.259--0.073) | 0.043<br>(-0.005-0.091) | -0.135<sup>++</sup><br>(-0.213--0.058) | -0.092<br>(-0.185-0) | -0.050<br>(-0.137-0.038) |
| **Level 3** | -0.270<sup>+++</sup><br>(-0.324--0.216) | -0.120<sup>++</sup><br>(-0.199--0.041) | -0.017<br>(-0.044-0.01) | -0.131<sup>+++</sup><br>(-0.194--0.067) | -0.220<sup>+++</sup><br>(-0.307--0.133) | -0.198<sup>+++</sup><br>(-0.275--0.122) |
| **Level 4** | -0.135<sup>+++</sup><br>(-0.166--0.103) | -0.134<sup>++</sup><br>(-0.214--0.055) | -0.042<sup>++</sup><br>(-0.069--0.015) | -0.186<sup>+++</sup><br>(-0.249--0.123) | -0.296<sup>+++</sup><br>(-0.383--0.209) | -0.174<sup>+++</sup><br>(-0.246--0.101) |

<sup>+++</sup>p<0.0001, <sup>++</sup>p<0.01, <sup>†</sup>p<0.05
The log beta values represent the log change in the risk ratio for each day a policy is in place. Effect sizes are from negative binomial models that adjust for: period; rural-urban continuum classification; 2018 county population percentage with at least a bachelor's degree, living in poverty, Black, and Hispanic; and 2018 net county migration rate.

Policy effects varied significantly across US Census regions. Intervention level 4 was the only policy to be associated with significant declines in COVID-19 case rates across all US Census regions, with a daily decline in the COVID-19 rate ranging from 25.7% (95% CI: 18.9-31.8%) in South Atlantic states to 4.1% (95% CI: 1.5-6.7%) in North Central States (Supplemental Figure B1, Supplemental Table B1).

Intervention level 3 significantly reduced COVID-19 rates in all regions except North Central States, while intervention level 2 reduced COVID-19 rates in Pacific, Mountain and South Central states. Intervention level 1 did not reduce COVID-19 in any US Census Region except South Atlantic states.

#### Doubling Time

Note that decreases in COVID-19 case rates are equivalent to increases COVID-19 doubling time. Thus, consistent with models for COVID-19 cases, intervention levels 3 and 4 significantly increased doubling time (log beta estimates 0.54 [95% CI: 0.22-0.86] and 0.45 [95% CI: 0.10–0.80], respectively, Table 2), while levels 1 and 2 did not (log beta estimates 0.20 [95% CI: –0.17-0.41] and 0.15 [95% CI: –0.17–0.46], respectively). Each day on intervention 1 reduced the log beta doubling time by 0.081 (95% CI: –0.12--0.04, p = 0.0005), while each day on intervention 4 increased log beta doubling time by 0.025 (95% CI: 0.01-0.04, p = 0.0017). The predicted marginal effects of interventions 3 and 4 on doubling time indicate that level 3 effects peak at approximately 25 days compared to 40 days for level 4 (Table 2, Figure 3).

**Table 2.** Predicted Doubling Time and 95% Confidence Intervals by Duration of each Intervention Level.

|  | Log Beta | (95% CI) |
| --- | --- | --- |
| **Fixed Intervention Effect:** |  |  |
| Level 1 | 0.107 | (-0.178-0.391) |
| Level 2 | 0.163 | (-0.153-0.48) |
| Level 3 | 0.565 | (0.251-0.878)<sup>++</sup> |
| Level 4 | 0.469 | (0.123-0.815)<sup>++</sup> |
| **Duration of Intervention Effect:** |  |  |
| Level 1 | -0.082 | (-0.127- -0.037)<sup>++</sup> |
| Level 2 | -0.036 | (-0.081-0.008) |
| Level 3 | -0.033 | (-0.055- -0.011)<sup>++</sup> |
| Level 4 | 0.012 | (-0.002-0.027) |

<sup>+++</sup>p<0.0001, <sup>++</sup>p<0.01, <sup>†</sup>p<0.05
The log beta values represent the log change in doubling time when an intervention policy is in place or the change for the number of days the policy is in effect.
Results are from a negative binomial model adjusted for the following: period; rural-urban continuum classification; population density; 2018 county level population percentage with at least a bachelor's degree, living in poverty, Black, and Hispanic; and net migration rate into the county in 2018.

**Figure 3.**
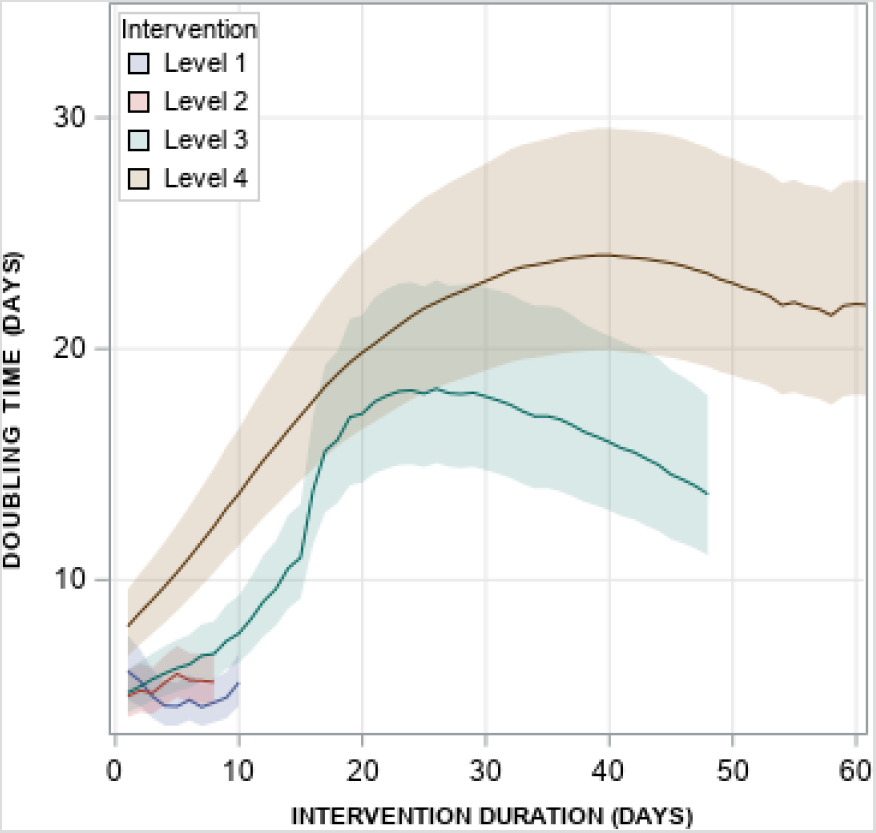
Predicted Doubling Times for each Intervention by Duration of Policy (with 95% Confidence Intervals) Intervention level effects are estimated using the full model results. Thus, each line represents the mean and 95% CI for all counties in the US through the 95^th^ percentile of duration (i.e., 48 days is the 95^th^ percentile of duration for policy level 3). Intervention levels 1 and 2 do not have observed data beyond 17 and 13 days, respectively.

Intervention effects on doubling time varied by US Census Region, but differently compared to the COVID-19 case rate models. Duration of intervention levels 3 and 4 were associated with increased doubling times only in Middle Atlantic and Pacific states, while level 2 achieves longer doubling time in Pacific states (Supplemental Table C1). Of concern, duration of level 3 policies was associated with lower doubling times in West South Central and New England states (log beta estimates –0.057 (95% CI –0.086- −0.028) and −0.033 (95% CI –0.055- −0.011), respectively), while duration of intervention levels 1 and 2 were associated with lower doubling times in West Central (North and South) and New England states (Supplemental Table C1). Predicted policy effects by policy duration for each region and the full multivariate model results for the doubling time model are presented in Supplemental Table C2 and Supplemental Figure C1.

#### Deaths

This analysis includes data from 73,676 COVID-19 deaths, the majority in Middle Atlantic states (36%), followed by East North Central (19%), New England (15%), South Atlantic (11%), and Pacific (7%) states. Only intervention level 4 was associated with reduced risk of death (Figure 4), with each day on intervention level 4 associated with an average 15% decline in the COVID-19 death rate, which ranged from a 25.5% decline in Middle Atlantic and New England states (log beta −0.294, 95% CI: −0.39-−0.19) to a 5.5% decline in North Central states (−0.056, 95% CI: –0.06-−0.05) (Figure 4, Supplemental Table D1). The COVID-19 death rate declined for intervention 3 only in Pacific states (log beta −0.146, 95% CI: −0.26-−0.03), while all other regions experienced an increase in death rates with increasing time on intervention level 3. Similarly, longer duration on intervention level 2 was associated with increases in death rates for all regions (Supplemental Table D1).

**Figure 4.**
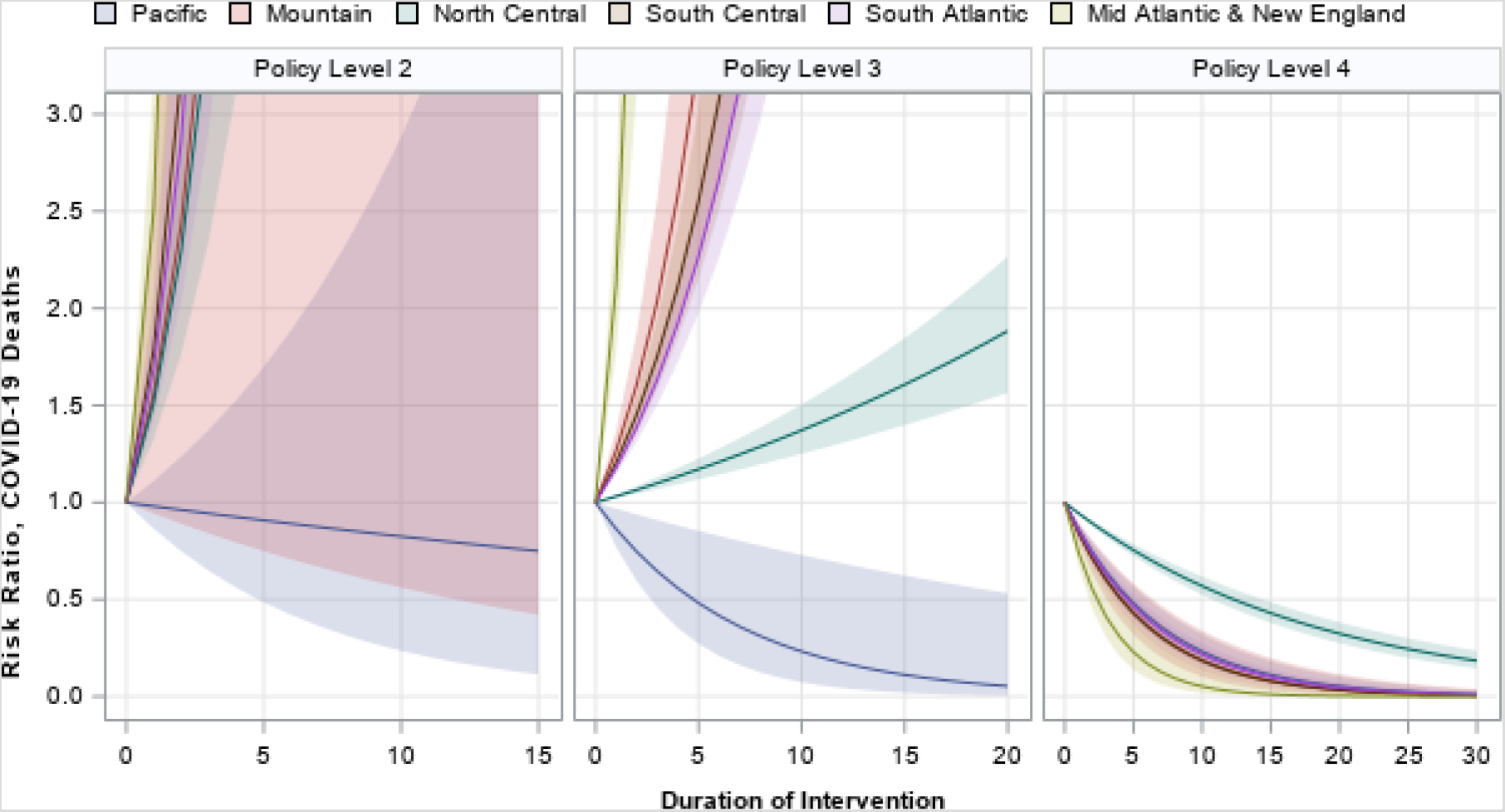
Risk Ratio and 95% Confidence Intervals of COVID-19 Deaths by Duration of Policy for each US Census Region.

### Socio-Environmental Disparities

In addition to policy intervention effects, COVID-19 cases and deaths were related to several socio-environmental factors in our models. Counties located in North Central, South Central and South Atlantic states had significant social and demographic associations with COVID-19 cases and deaths that were not as prevalent in other US regions (Supplemental Table B1). Notably, these three regions had significant positive relationships between Black and Hispanic population size and rates of COVID-19 cases, but also lower COVID-19 case rates in counties with higher percentages of people living in poverty. These three regions, plus Middle Atlantic states, are the only regions with significant correlations between percentages of the county population living in poverty and the percentage of the population Black (Pearson correlations > 0.22 for all regions, data not shown). COVID-19 case rates (and doubling time) were highest (lowest doubling times) in metropolitan counties or in counties adjacent to metropolitan areas, and lowest (highest doubling times) in rural counties and counties with under 20,000 population.

The relationship between climate parameters and COVID-19 was also variable, and these relationships often differed between case doubling time and deaths (Supplemental Figure D1, D2). Solar radiation, for example, which is frequently invoked as a negative forcing on COVID-19 transmission on account of its relationship with UV radiation intensity [33, 34], is associated with increased case doubling time (i.e., decreased transmission) in eastern regions but with decreased doubling time in western regions. For deaths, most regions tend towards decreased deaths with increased solar radiation (significantly so in the Pacific, West North Central, West South Central, and Middle Atlantic regions), but this is not true in all regions or for the contiguous United States as a whole.

Increases in specific humidity and in minimum temperature were associated with decreased doubling time and increased deaths for the county as a whole (specific humidity: log beta −0.046, 95% CI −0.07-−0.027 for doubling time, 0.052, 95% CI 0.005-0.10 for deaths; minimum temperature: not significant for doubling time, log beta 0.045, 95% CI 0.025–0.065 for deaths). However, these results differed by region and by response variable: the Pacific region, showed significant declines in death rates with increased specific humidity and minimum temperature, but also decreased doubling time for increases in both variables (significantly for specific humidity). New England, in contrast, showed significant increases in deaths and decreases in doubling time with specific humidity, and most regions had no statistically significant relationship. Once again, analyses of case doubling time were not always consistent with analyses of death rates.

## Discussion

During the period at which COVID-19 emerged until the initiation of phased reopening of US state businesses and public places, NPIs that include non-essential business closures, suspending non-violent arrests, suspending elective medical procedures, suspending evictions, restricting mass gatherings of at least 10 people, sheltering in place / stay-at-home, public mask requirements, and travel restrictions, are the most effective at slowing COVID-19 propagation as measured by the rate of new cases detected and doubling time. Further, our models demonstrate that only the strictest interventions—sheltering in place, public mask requirements and travel restrictions—have been effective at reducing COVID-19 death rates across all US Census Regions.

The results of this study are consistent with recent findings from Wuhan [35] and Europe [36] that reported significant declines in the effective reproductive number following implementation of NPIs that included quarantine, travel restriction, shelter-in-place, school and business closures, and social distancing. Results are also consistent with studies in the US showing the effect of shelter-in-place along the Iowa-Illinois border [5] and a meta-analysis of shelter-in-place effects on COVID-19 infection [37]. This analysis furthers our knowledge by evaluating the complete set of NPIs, including timing of their implementation and duration, and how their effects differed across US regions. Further, after adjusting for all known covariates, we find that the peak intervention effects on doubling time occurred approximately 25 and 40 days after intervention levels 3 and 4 were implemented, respectively. Why the effects leveled off yet the models indicate the NPIs should maintain a decline in new case rates (and deaths) remains an important area of research. It is likely that issues related to NPI adherence, transportation, and racial and economic disparities play a role in the long-term effectiveness of each NPI level. The timing of policy implementation may also introduce heterogeneity. We observed that states experiencing early cases (Pacific, New England, and Mid-Atlantic) had significantly longer gap times between case detection and NPI initiation than states experiencing their first COVID-19 case later. This difference in policy initiation time is likely due to non-coastal US Regions learning from experience of coastal regions to implement NPIs more quickly. As states continue to remove NPI protections, results from this study help inform areas of research to improve policy decision-making to reduce COVID-19 case and death rates.

In addition to evaluating policy effects, this study reported the effects of race, poverty and climate parameters related to COVID-19. Consistent with previously published data, we found that counties with higher percentages of Black and Hispanic populations tend to report more cases. However, our analysis suggests that this relationship primarily exists in Southern and Central US states. In addition, we have mixed results for climate predictors of COVID-19 that inform related environmental research, including: the instability of climate variables at this stage of the epidemic, particularly with choice of response variable; the inter-regional variability in climate sensitivities that indicate large scale analyses are not necessarily representative of regional climate influence; and the difficulty in isolating climate effects from policy and socio-demographic factors. We recommend further research be conducted at different spatial scales (community, census tract, etc.) to better characterize the climate-COVID-19 relationship.

There are several limitations to note in our study. First, the analysis does not have an accurate representation of the availability of testing (or the number of tests administered) at the county level for the time series. As this availability changed over time for all counties in the US, we cannot accurately characterize the population at risk for COVID-19 case detection. However, we are confident in COVID-related deaths reported. Second, we do not have measures of public acceptance and adherence to NPIs. Anecdotal data suggest significant variation of NPI adherence and that with phased reopening of some states, NPIs were viewed as an affront to civil liberties. Data on NPI adherence are critical to accurately measuring “dose” of the NPI to accurately characterize the population-level effect. Third, we do not look at individual interventions. However, this is a choice rather than limitation as sets of NPIs are considered a more appropriate response than single interventions, which have never been employed historically without others. Finally, we would ideally be applying these methods to a randomized design, which is impossible for COVID-19. Thus, our inferences are drawn from observational data.

## Conclusion

The most aggressive NPIs (shelter-in-place, public mask requirements, and travel restrictions) were the only policies that significantly decreased both COVID-19 cases and deaths in the US between January 2020 and the phased re-opening of states. Socio-environmental factors, including racial/ethnic status and specific humidity, potentially contribute to heterogeneity of COVID-19 propagation and NPI mitigation effects. These results may inform public health policy as states continue to cautiously reopen their economies.

## Data Availability

All data are available on public databases.

https://coronavirus.jhu.edu/

https://data.census.gov/mdat/#/

https://www.nga.org/coronavirus/#resElections

https://www.multistate.us

https://www.ers.usda.gov/data-products/rural-urban-continuum-codes/

